# Predicting mortality in SARS-COV-2 (COVID-19) positive patients in the inpatient setting using a Novel Deep Neural Network

**DOI:** 10.1101/2020.12.13.20247254

**Authors:** Maleeha Naseem, Hajra Arshad, Syeda Amrah Hashmi, Furqan Irfan, Fahad Shabbir Ahmed

## Abstract

**Background:** The second wave of COVID-19 pandemic is anticipated to be worse than the initial one and will strain the healthcare systems even more during the winter months. Our aim was to develop a machine learning-based model to predict mortality using the deep learning Neo-V framework. We hypothesized this novel machine learning approach could be applied to COVID-19 patients to predict mortality successfully with high accuracy.

**Methods:** The current Deep-Neo-V model is built on our previously statistically rigorous machine learning framework [Fahad-Liaqat-Ahmad Intensive Machine (FLAIM) framework] that evaluated statistically significant risk factors, generated new combined variables and then supply these risk factors to deep neural network to predict mortality in RT-PCR positive COVID-19 patients in the inpatient setting. We analyzed adult patients (≥18 years) admitted to the Aga Khan University Hospital, Pakistan with a working diagnosis of COVID-19 infection (n=1228). We excluded patients that were negative on COVID-19 on RT-PCR, had incomplete or missing health records. The first phase selection of risk factor was done using Cox-regression univariate and multivariate analyses. In the second phase, we generated new variables and tested those statistically significant for mortality and in the third and final phase we applied deep neural networks and other traditional machine learning models like Decision Tree Model, k-nearest neighbor models and others.

**Results:** A total of 1228 cases were diagnosed as COVID-19 infection, we excluded 14 patients after the exclusion criteria and (n=)1214 patients were analyzed. We observed that several clinical and laboratory-based variables were statistically significant for both univariate and multivariate analyses while others were not. With most significant being septic shock (hazard ratio [HR], 4.30; 95% confidence interval [CI], 2.91-6.37), supportive treatment (HR, 3.51; 95% CI, 2.01-6.14), abnormal international normalized ratio (INR) (HR, 3.24; 95% CI, 2.28-4.63), admission to the intensive care unit (ICU) (HR, 3.24; 95% CI, 2.22-4.74), treatment with invasive ventilation (HR, 3.21; 95% CI, 2.15-4.79) and laboratory lymphocytic derangement (HR, 2.79; 95% CI, 1.6-4.86). Machine learning results showed our DNN (Neo-V) model outperformed all conventional machine learning models with test set accuracy of 99.53%, sensitivity of 89.87%, and specificity of 95.63%; positive predictive value, 50.00%; negative predictive value, 91.05%; and area under the curve of the receiver-operator curve of 88.5.

**Conclusion:** Our novel Deep-Neo-V model outperformed all other machine learning models. The model is easy to implement, user friendly and with high accuracy.

## INTRODUCTION

Severe acute respiratory syndrome coronavirus 2 (SARS-COV 2) has caused 60 million infections and 1.4 million deaths worldwide (1) and 800, 000 deaths in the United States (2). Despite strict measures being deployed and special instructions given to the mass public, the second wave is anticipated to be far worse than the first one (3). Some countries have started to ease those earlier restrictions because of economic implications from the initial lockdown, which may create a further deepening of the current crisis, as cases continue to rise. This could overwhelm the already strained healthcare systems across the United States and the world.

Machine learning has been extensively used in the automotive, defense and fin-tech industry over the past couple of years with great success. The use of these systems to predict health outcomes have been limited. Epidemic Renormalization Group (eRG) has used a machine learning framework to predict the time evolution of the first and second wave based on the data from the first wave in Europe (4). We have previously developed algorithms that predict mortality in the clinical setting and performed better than most clinical scales utilized currently to predict mortality (5-7) During the current COVID-19 pandemic crisis, the aim was to develop a mortality prediction tool that can predict death in COVID-19 patients at admission. This would help the already strained healthcare systems and physicians around the world in crucial clinical decision making, resource management and family-counselling. We hypothesize that machine learning, specifically deep-learning could be applied to COVID-19 patients with high accuracy. Using deep-learning to predict mortality in these patients may assist in clinical decision making, risk stratification and planning strategies in future for such pandemics at a larger scale. Not much work has been done in mortality prediction in COVID-19 patients in with lower socio-economic countries (LMIC) or developed countries. Our hypothesis is that machine learning could be applied to COVID-19 patients with high accuracy. Hence, predicting mortality and clinical outcomes using ML algorithms may assist in clinical decision making, risk stratification and planning strategies in future for such pandemics at a larger scale.

## METHODS

### Clinical setting and dataset

Clinical data was conducted at the Aga Khan University Hospital (AKUH). All patients’ records were completely anonymous, and the data collected has received Institutional Review Board/Ethical Review Committee (IRB/ERC) approval from Aga Khan University Hospital (AKUH), Pakistan. The dataset was de-identified and our study complied with the ethical principles recommended by Helsinki declaration (1964) and its amendments. We retrospectively collected data from AKUH – electronic medical record (EMR) that were admitted with a primary diagnosis of COVID-19 infection to the hospital between February 2020 and September 2020.

### Patient data collection and selection criteria

We included adult patients (>18 years of age) that were admitted to the hospital with a diagnosis of COVID-19 or were tested positive during their admission on reverse_transcriptase polymerase chain reaction (RT-PCR) based on Center for Disease Control and Prevention (CDC) and College of American Pathologist (CAP) guidelines (8, 9). Data was collected on demographics, and comorbidities at admission, the first 24-hours of laboratory investigations (hematological and blood biochemistry, (**Table 1**.), imaging and complete clinical characteristics (history, examination, treatment, hospital course and outcomes). We excluded all patients that had RT-PCR negative tests for COVID-19 and incomplete records or inaccurate medical record information.

**Table 1.**
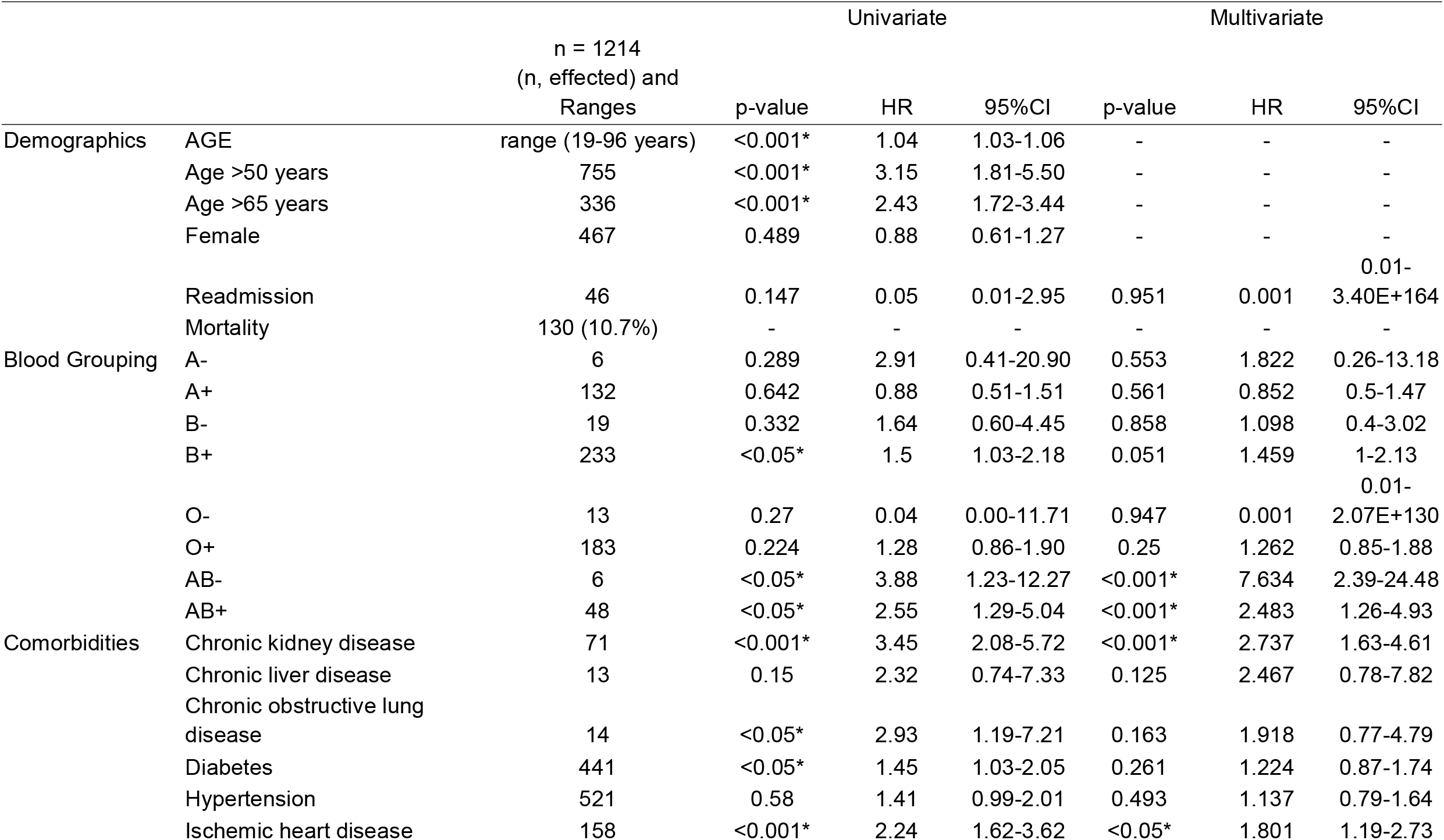
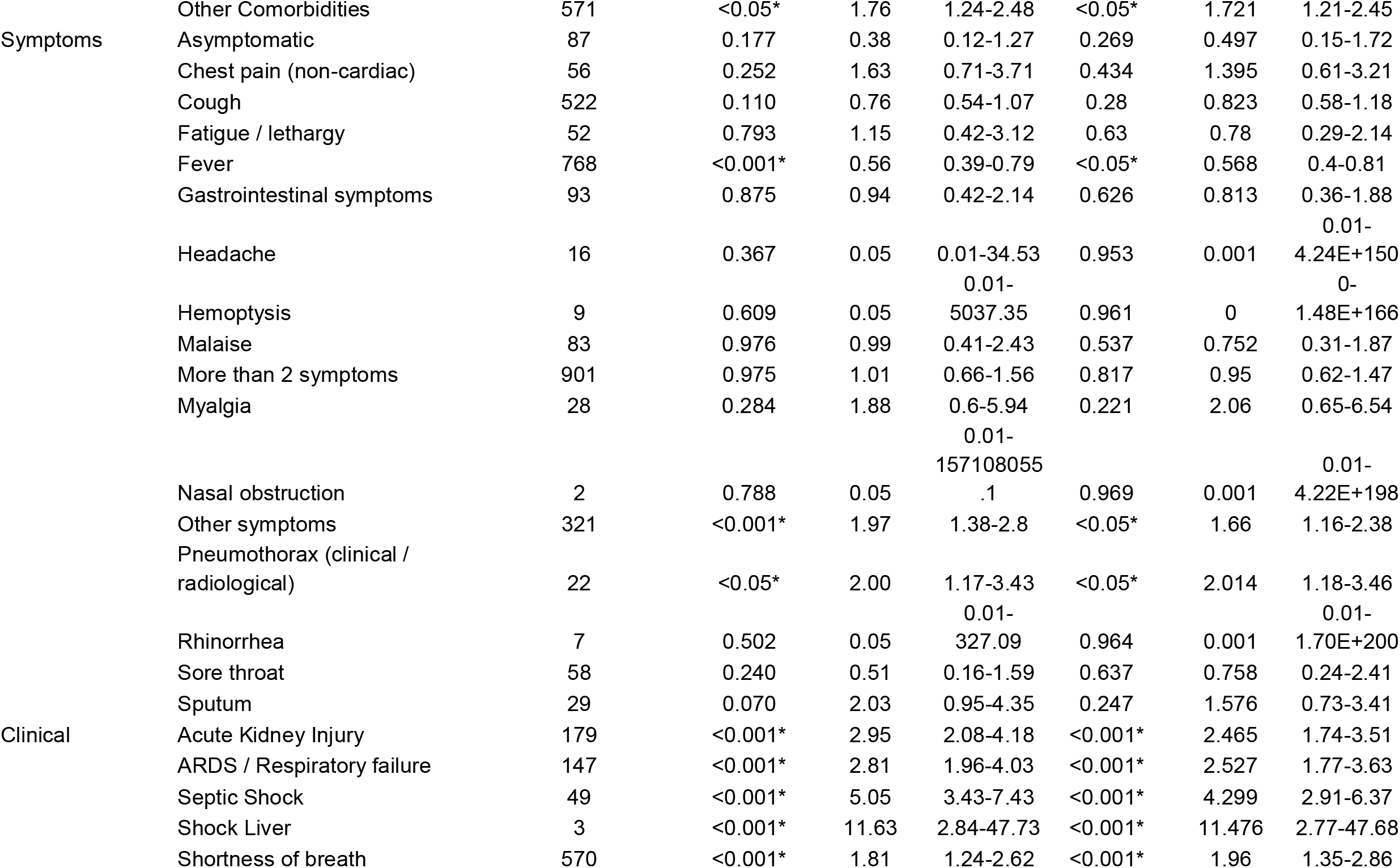
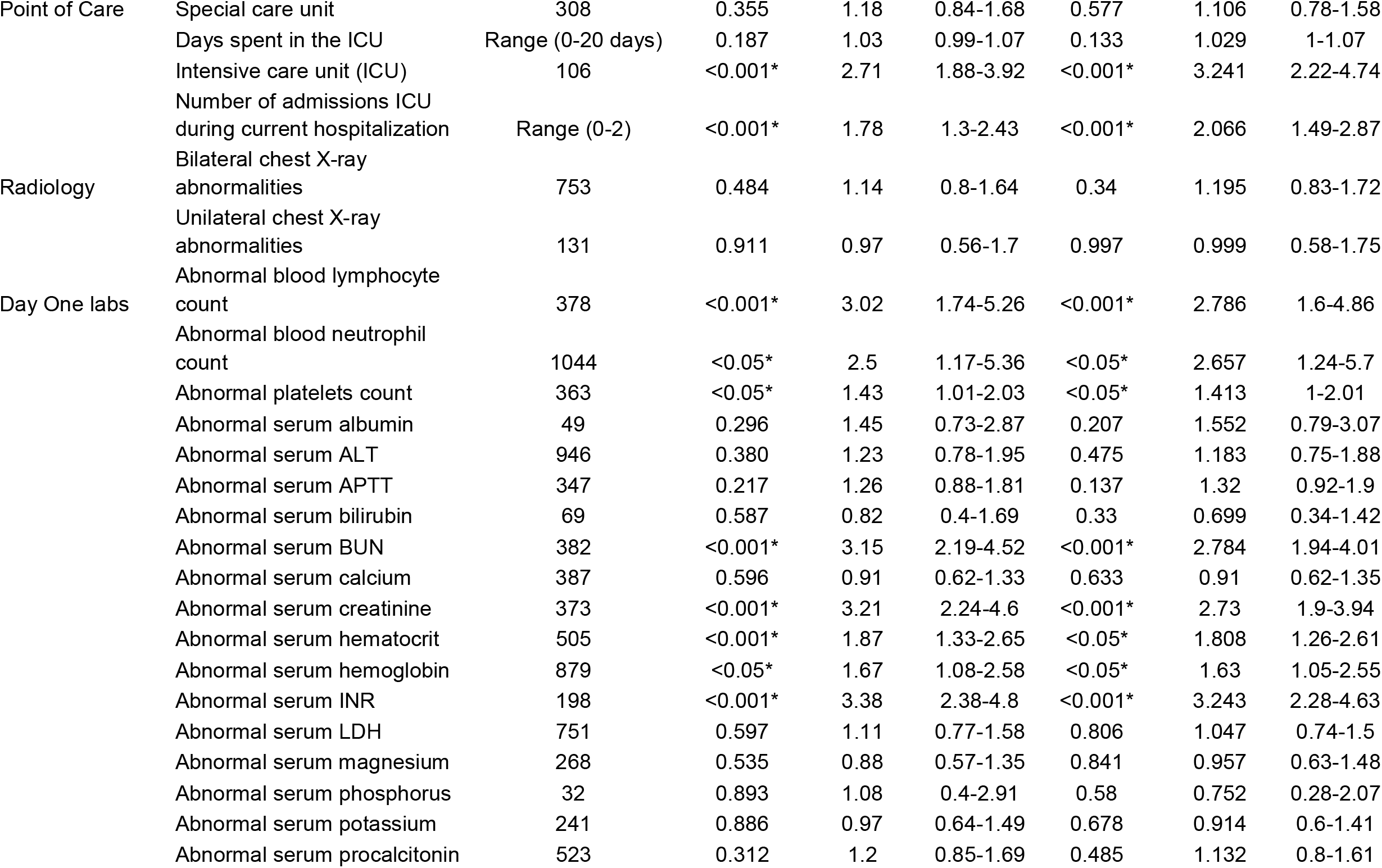
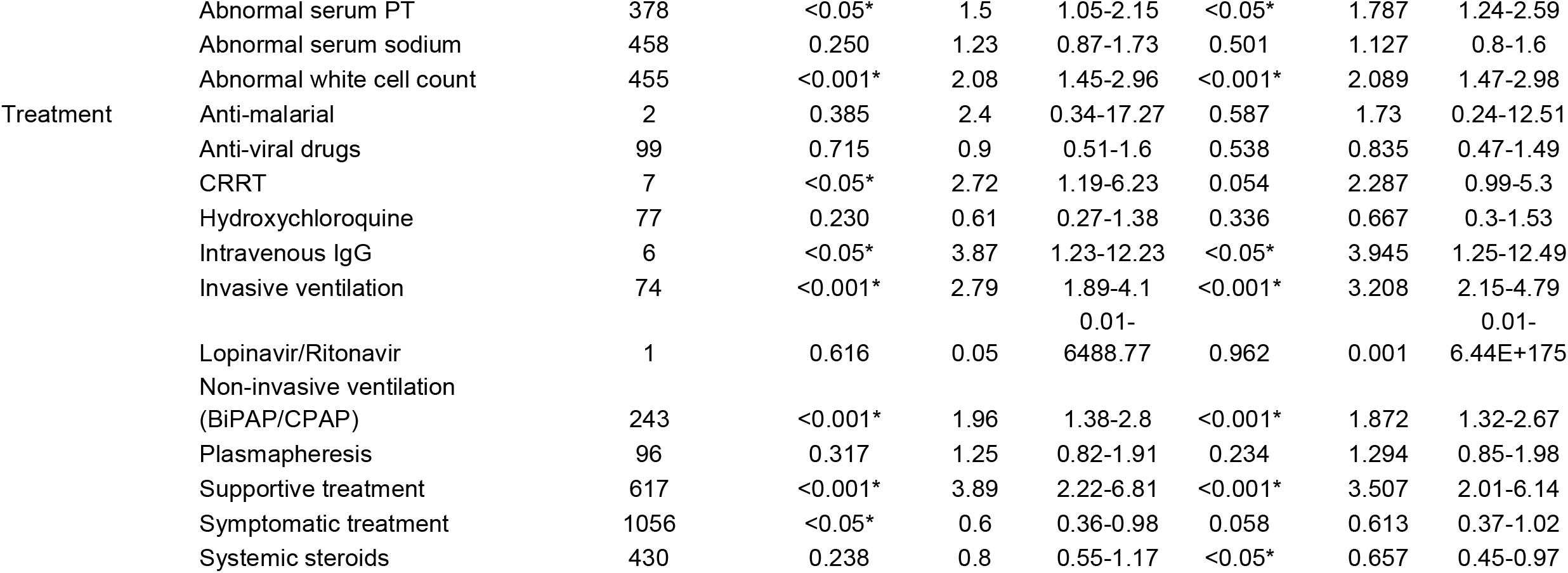
Demographics with Univariate and multivariate analysis of clinical variables as part of Phase I of the Neo-V and FLAIM machine learning frameworks.

### The Neo-V Framework

Neo-V is a tri-phase bio-statically rigorous machine learning that builds on our previous framework that had better accuracy then the currently used clinical scoring systems in predicting mortality in the intensive care unit (ICU) patients(5, 6).

**Phase I:** Also known as the statistical-phase; in which data was analyzed by univariate and multivariate Cox-regression analysis (X) using IBM SPSS (version 24.0.0.0) (X) for outcome assessment with hazard ratio and confidence intervals. A *p*-value of <0.05 was considered statistically significant. We also did demographic data frequency analysis. Statistical analysis was carried out on all the variables included in **Table 1**.

**Phase II:** In contrast to our previously published model we created new variables for the existing dataset called neo-variables. These variables included a combination of two clinically relevant labs that were significant in both the univariate and the multivariate analysis **(Table 1.)**. These variables also underwent univariate and multivariate analysis for outcome assessment with hazard ratio and confidence intervals

**Phase III:** Biological datasets are highly imbalanced with respect to the outcomes (i.e. more people were survived, then those who didn’t) and machine learning models are very sensitive to imbalanced data and can produce variable and non-reproducible results. To address this, we optimized the dataset using Synthetic Minority Over-sampling Technique (SMOTE) algorithm during the training process (10). In the machine learning phase, we used all variables that were statistically significant in phase I and II in both the univariate and multivariate analysis (non-significant risk factors were excluded). The final dataset was randomized and divided into a training and testing set with a 70/30 percent split respectively (30% data left out to test the models). After partitioning the data, we allocated feature vectors of the training instances by X_train with corresponding outcome label as Y_train. Similarly, for the test set we allocated X_test and Y_test as testing vector instances and corresponding outcomes, respectively. The models trained on X_train and Y_train. The models tried to learn the behavior/distribution of the data and generate a hypothesis/fitting function. Once the training is concluded the model will then test the X_test and produce an output (prediction) called Y_pred. A comparison is done between Y_pred and Y_test. We had previously discussed that reduction of the number of irrelevant risk factors can produce better performances and significantly improve classifications. In this study we used conventional machine learning models including Random Trees (CART), K-Nearest Neighbor (KNN), Support Vector Classifier - Radial Basis Function (SVC - RBF), Ada-Boost-Classifier (ABC) and Quadratic Discriminant Analysis (QDA) and a deep neural network (DNN).

### Deep-FLAIM

FLAIM framework only has phase I and III and we used it to compare it with Neo-V Framework. The Deep-FLAIM model is a 4 layered model and details have been reported previously(5).

### Performance, Primary and Secondary Outcomes Analyses

Performance of all models was evaluated by comparing their accuracies and area under the receiver-operator curves (AUROC). Primary outcomes included sensitivity and specificity, while secondary outcomes included positive predictive values (PPV) and negative predictive values (NPV).

## RESULTS

From a total of 1228 patients we selected 1214 patients that were adult patients with complete data and RT-PCR proven COVID-19 infections. Demographics of this population showed a median age of 55 years (range 19-96 years), around 28% (n=336) of the admitted population being elderly (>65 years of age). Median length of stay 5 days (range 1-54 days), most patients admissions were male (61.5%).

The clinical characteristics of these patients included hypertension (43%, n=521) and diabetes (36%, n=441) being the most common comorbid. Presenting symptoms ranged widely from being asymptomatic to shortness. The most significant clinical risk factors for death during the hospital admission included chronic kidney disease (n=71, HR=2.74, 95%CI=1.63-4.61), ischemic heart disease (n=158, HR=1.80, 95%CI=1.19-2.73), other comorbidities (n=571, HR=1.72, 95%CI=1.21-2.45), shortness of breath (n=570, HR=1.96, 95%CI=1.35-2.86), other symptoms (non-respiratory and non-gastrointestinal symptoms, n=321, HR=1.66, 95%CI=1.16-2.38), acute kidney injury (n=179, HR=2.47, 95%CI=1.74-3.51), acute respiratory distress syndrome / respiratory failure (n=147, HR=2.53, 95%CI=1.77-3.63), septic shock (n=49, HR=4.30, 95%CI=2.91-6.37), intensive care unit (ICU) admission (n=106, HR=3.24, 95%CI=2.22-4.74), number of ICU admissions during current hospitalization (range=0-2, HR=2.01, 95%CI=1.49-2.87), invasive ventilation (n=74, HR=3.21, 95%CI=2.15-4.79), non-invasive ventilation (BiPAP/CPAP) (n=243, HR=1.87, 95%CI=1.32-2.67), supportive treatment (n=617, HR=1.87, 95%CI=1.32-2.67) and blood group AB+ (n=48, HR=2.48, 95%CI=1.26-4.93); while the utilization of systemic steroids (n=430, HR=0.66, 95%CI=0.45-0.97) and presence of fever (n=768, HR=0.57, 95%CI=0.40-0.81) were associated with better overall all survival. The most significant (statistical association with mortality) laboratory abnormalities for these patients included white cell count (n=455, HR=2.09, 95%CI=1.47-2.59), lymphocyte counts(n=378, HR=2.79, 95%CI=1.60-4.86), neutrophil count (n=1044, HR=2.67, 95%CI=1.24-5.70), platelets count (n=363, HR=1.41, 95%CI=1.00-2.01), hematocrit (n=505, HR=1.81, 95%CI=1.26-2.61), hemoglobin (n=879, HR=1.63, 95%CI=1.05-2.55), blood urea nitrogen (BUN, n=382, HR=2.78, 95%CI=1.94-4.01), creatinine (n=373, HR=2.73, 95%CI=1.90-3.94), international normalized ratio (INR, n=198, HR=3.24, 95%CI=2.28-4.63), prothrombin time (PT, n=378, HR=1.79, 95%CI=1.24-2.59). There were some risk factors that were significant for mortality but had too few patients including; shock liver (n=3, HR=11.47, 95%CI=2.77-47.68), blood group AB- (n=6, HR=7.63, 95%CI=2.39-24.48), Rhinorrhea (n=7, HR=0.001, 95%CI=0.01-1.7e200), treatment with intravenous IgG (n=6, HR=3.95, 95%CI=1.25-12.49) and pneumothorax (clinical or radiological, n=22, HR=2.01, 95%CI=1.18-3.46). **Table 1**. shows results from univariate and multivariate analysis (hazard ratios, confidence intervals and *p*-values) of all the clinical and laboratory data.

In phase II (new variable phase) we created 11 new variables that included; total number of comorbidities (range 0-6, HR=1.30, 95%CI=1.14-1.49), more than 2 comorbidities (n=499, HR=1.79, 95%CI=1.23-2.61), total number of symptoms(range 0-6, HR=1.07, 95%CI=0.91-1.26), total types of treatments received (range 0-6, HR=1.27, 95%CI=1.11-1.46), abnormal labs: creatinine or blood urea nitrogen (CR or BUN, (n=517, HR=0.36, 95%CI=0.36-0.89)), hemoglobin or hematocrit (HB or HCT, (n=883, HR=0.56, 95%CI=0.36-0.89)), platelets or INR (PLT or INR, (n=460, HR=2.09, 95%CI=1.47-2.98)), PT or INR (PT or INR, (n=502, HR=0.414, 95%CI=)), total leukocyte counts of lymphocytes (TLC or LYMP, (n=0.19, HR=, 95%CI=0.09-0.43)), total leukocyte counts or neutrophils (TLC or NEU, (n=1085, HR=0.18, 95%CI=0.06-0.57)), total number of abnormal labs(range 0-17, HR=1.22, 95%CI=1.15-1.30), univariate and multivariate statistical analysis can be seen in **Table 2**. The new variables in phase II referred to as ‘neo-variables’ were statistically significant in the uni and the multivariate analysis except the total number of symptoms.

**Table 2.**
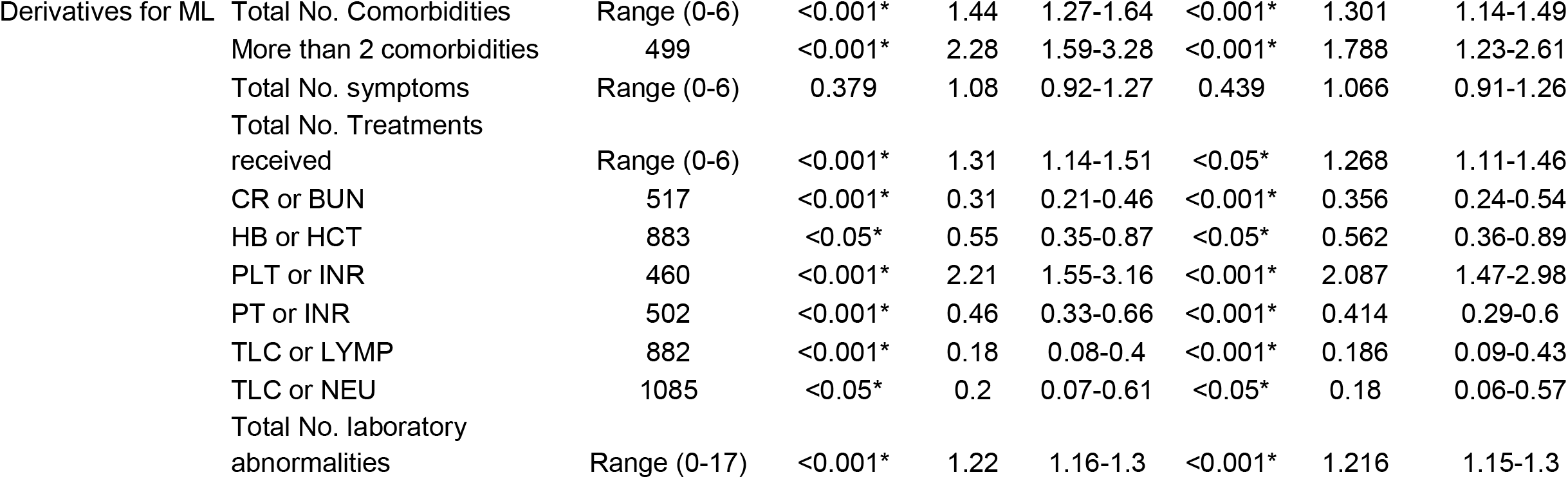
Derivative variables and there univariate and multivariate analysis with mortality during hospital stay.

|  |  |  |  |  |  |  |  |  |
| --- | --- | --- | --- | --- | --- | --- | --- | --- |
| Derivatives for ML | Total No. Comorbidities | Range (0-6) | <0.001* | 1.44 | 1.27-1.64 | <0.001* | 1.301 | 1.14-1.49 |
|  | More than 2 comorbidities | 499 | <0.001* | 2.28 | 1.59-3.28 | <0.001* | 1.788 | 1.23-2.61 |
|  | Total No. symptoms | Range (0-6) | 0.379 | 1.08 | 0.92-1.27 | 0.439 | 1.066 | 0.91-1.26 |
|  | Total No. Treatments received | Range (0-6) | <0.001* | 1.31 | 1.14-1.51 | <0.05* | 1.268 | 1.11-1.46 |
|  | CR or BUN | 517 | <0.001* | 0.31 | 0.21-0.46 | <0.001* | 0.356 | 0.24-0.54 |
|  | HB or HCT | 883 | <0.05* | 0.55 | 0.35-0.87 | <0.05* | 0.562 | 0.36-0.89 |
|  | PLT or INR | 460 | <0.001* | 2.21 | 1.55-3.16 | <0.001* | 2.087 | 1.47-2.98 |
|  | PT or INR | 502 | <0.001* | 0.46 | 0.33-0.66 | <0.001* | 0.414 | 0.29-0.6 |
|  | TLC or LYMP | 882 | <0.001* | 0.18 | 0.08-0.4 | <0.001* | 0.186 | 0.09-0.43 |
|  | TLC or NEU | 1085 | <0.05* | 0.2 | 0.07-0.61 | <0.05* | 0.18 | 0.06-0.57 |
|  | Total No. laboratory abnormalities | Range (0-17) | <0.001* | 1.22 | 1.16-1.3 | <0.001* | 1.216 | 1.15-1.3 |

The performance of our previously designed Deep-FLAIM model was compared to the Neo-V framework (including Deep-Neo-V and other conventional machine learning algorithms) see **Table 3**. Performance results show Deep-FLAIM (training accuracy = 86.7%, testing accuracy = 84.7%, sensitivity = 68.9, specificity = 86.9, PPV = 42.5, NPV = 95.2, FPR =13.1 and AUROC = 86.9); while conventional machine learning show: Random Forest (RF, training accuracy = 94.9%, testing accuracy = 85.8%, sensitivity = 28.9, specificity = 93.4, PPV = 32.4, NPV = 90.4, FPR = 6.25 and AUROC = 69.5), k-Nearest Neighbors (k-NN, training accuracy = 92.3%, testing accuracy = 77.8%, sensitivity = 42.2, specificity = 82.8, PPV = 25.7, NPV = 91.1, FPR = 17.2 and AUROC = 66.5), Simple Vector Classifier – Radial Basis Function (SVC – RBF, training accuracy = 69.0%, testing accuracy = 65.8%, sensitivity = 75.6, specificity = 64.4, PPV = 23.0, NPV = 94.9, FPR = 35.6 and AUROC = 80.8), Decision Trees (DT, training accuracy = 98.9%, testing accuracy = 85.2%, sensitivity = 42.2, specificity = 91.3, PPV = 40.4, NPV = 91.8, FPR = 8.8 and AUROC = 66.7), Adaptive Boosted Classifier (ABC, training accuracy = 90.1%, testing accuracy = 85.5%, sensitivity = 48.9, specificity = 90.6, PPV = 42.3, NPV = 82.7, FPR = 9.4 and AUROC = 77.65) and Quadratic discriminant analysis (QDA, training accuracy = 94.6%, testing accuracy = 87.4%, sensitivity = 60.0, specificity = 91.3, PPV = 49.1, NPV = 94.2, FPR = 8.74 and AUROC = 80.84). While our best model was a deep neural network (Deep-Neo-V) with training accuracy = 98.7%, testing accuracy = 87.7%, sensitivity = 33.3, specificity = 95.3, PPV = 50.0, NPV = 91.1, FPR = 4.69 and AUROC = 88.5. All receiver operator curves are shown in **Supplemental Figure X**.

**Table 3.**
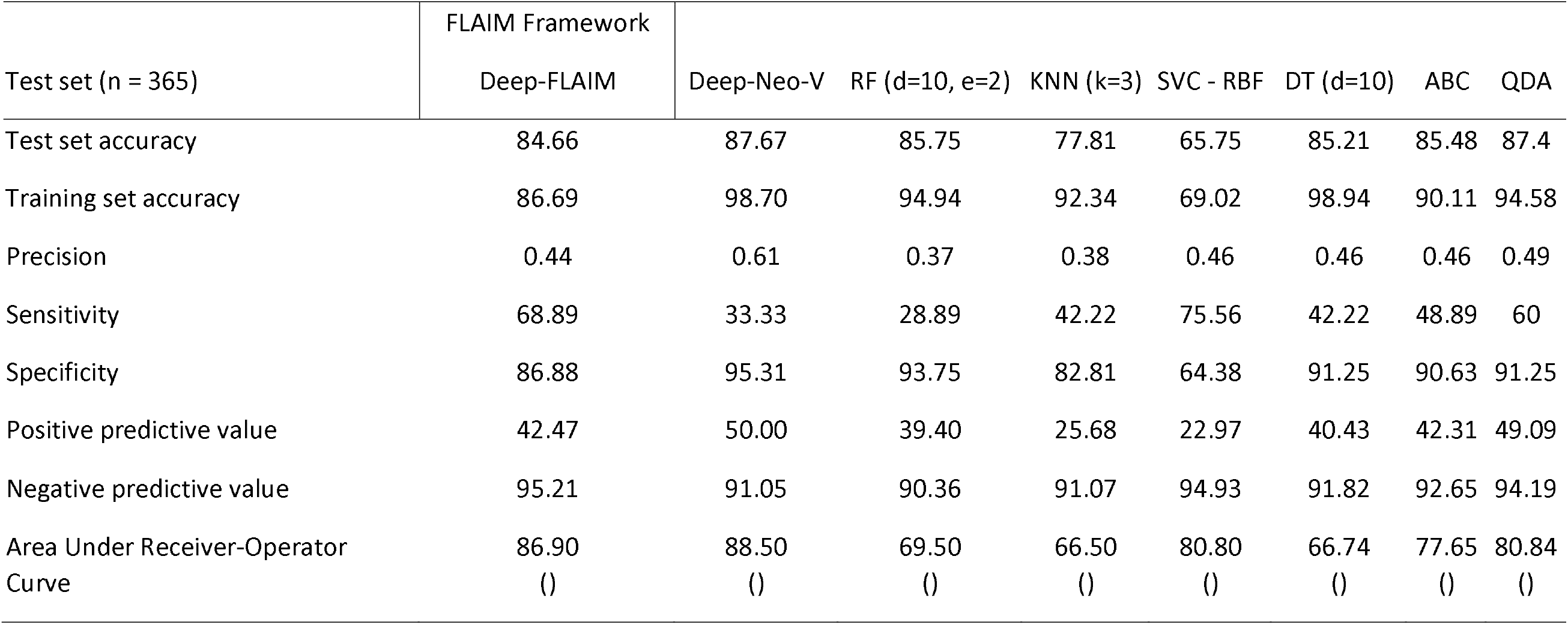
Primary and secondary outcomes machine learning results. Test set accuracy, training set accuracy, precision, sensitivity, specificity, positive predictive value, negative predictive value and AUROC.

**Table 4.** Comparison of currently available Deep learning models for mortality in the inpatient setting.

|  | Current | Li X et al, 2020 |
| --- | --- | --- |
| Sample Size (n) | 1214 | 1108 |
| Clinical setting | In-patient | In-patient |
| Training and Test (set) split | 70/30 | 90/10 |
| Datapoint / Variables | 47 | 15 |
| Testing accuracy | 87.67 | 85.3 |
| Training accuracy | 98.70 | 89.2 |
| AUROC | 0.885 | 0.844 |
| Sensitivity | 33.33 | 75 |
| Specificity | 95.31 | 87.2 |
| PPV | 50.00 | 52.2 |
| NPV | 91.05 | 94.9 |

## DISCUSSION

As the second wave of COVID-19 has started to unfold the already stained healthcare systems globally are being pushed to the limit with hospital and intensive care unit (ICU) beds reaching full-capacity. Impact of the virus has been global with developed countries even struggling with infection rates and hospitalization (11). The second wave is anticipated to be much tougher than the first one (12). With new vaccines on the horizon infection rates in the United states have skyrocketed to 1.34 million cases being diagnosed in the second week of December 2020 and more than 1100 deaths (13). The vaccines have just received FDA approval for widespread use of the vaccine for prevention of the disease while the actual logistics and distribution plans are still unknown (14). However, there is a need for the existence of clinical biomarkers and predictive models for mortality in these patients. machine learning has been used to predict mortality in cancer(15), cardiac disease (16); while our own work on mortality prediction has been on trauma patients, postoperative ileus cases in the ICU (5, 6) and diverticulitis in the inpatient setting (17) with good accuracies. A lot of epidemiological studies reporting clinical, laboratory and mortality outcomes have been done worldwide including developed and developing countries, but very few actually reported or developed a machine learning model for predicting the outcomes with a set accuracy.

In the current study we developed a new method of machine learning (Neo-V Framework) that uses a smaller amount of cases to train a deep neural network to give better predictions. This model is different from our previous FLAIM Framework (two-phase) and has a tri-phase structure (**Figure 1**.) Clinically we used just 1214 hospitalized patients for mortality prediction in RT-PCR positive COVID-19 cases using only data from the first 24-hours after admission. Clinical data analysis showed that with increasing age the patients’ mortality also increases. There are number of clinical risk factors that were associated with worse outcomes and documented in the clinical literature like chronic obstructive lung disease [COPD](18), chronic kidney disease [CKD](19), ischemic heart disease [IHD](20), pneumothorax (radiological or clinical diagnosis)(21), acute respiratory syndrome [ARDS](22), septic shock(23), shortness of breath(24), ICU admission(25), AB+ Blood group(26) and recurrent admission to the ICU. Hematological labs that were associated with mortality (previously presented in the results) were also seen in other studies(27). Biochemical laboratory abnormalities like creatinine, blood urea nitrogen(28), INR and PT(29) were also associated with mortality. Patients managed with invasive ventilation(30) and non-invasive ventilation (31) which actually signifies that the patients that were not able to maintain normal respiratory physiology had worse outcomes. Having fever(32) and the use of systemic steroids(33) as an early symptom of COVID-19 had better prognosis in our patient population.

**Figure 1.**
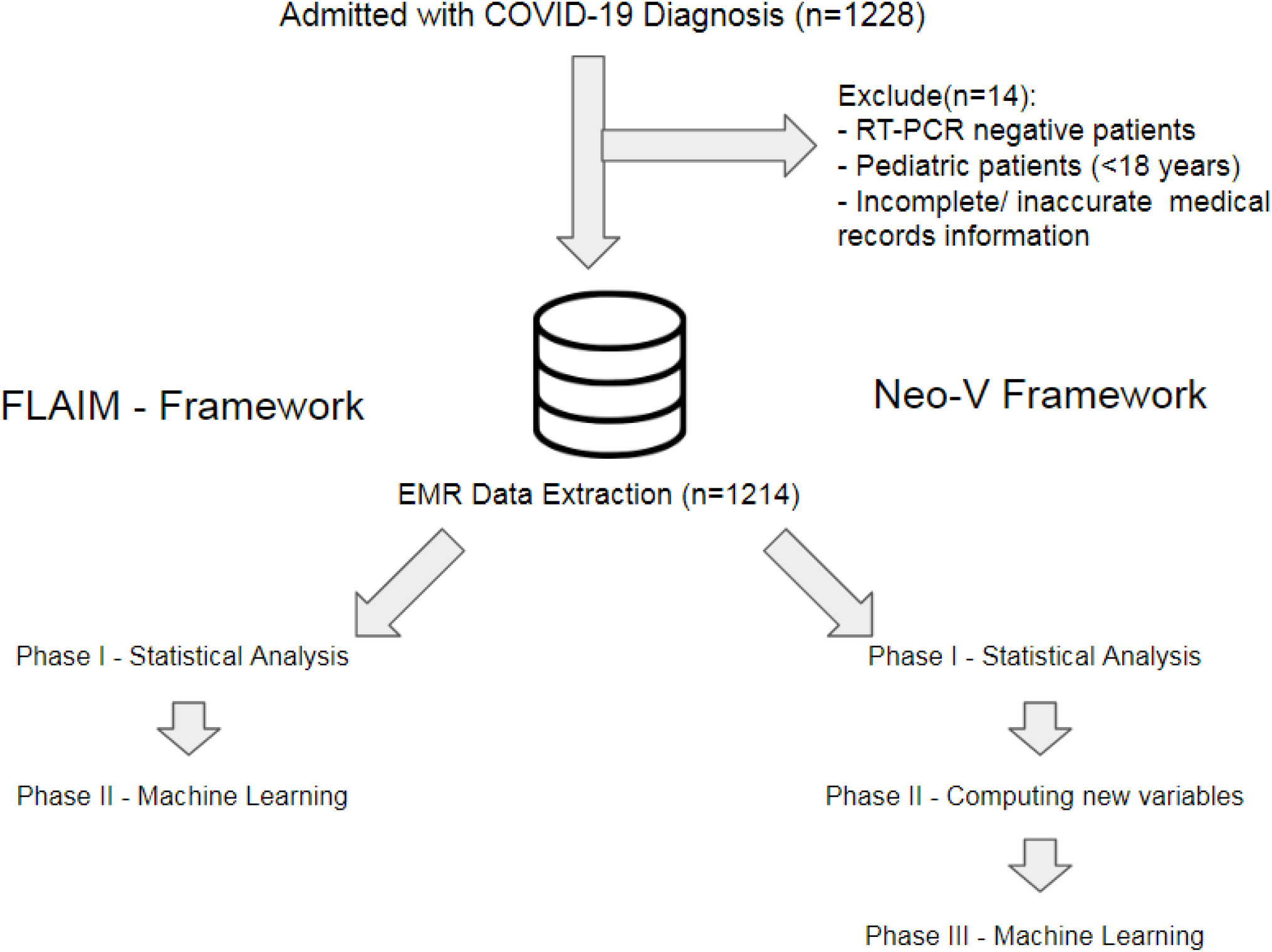
Experimental design of the study and the machine frameworks

**Figure 2.**
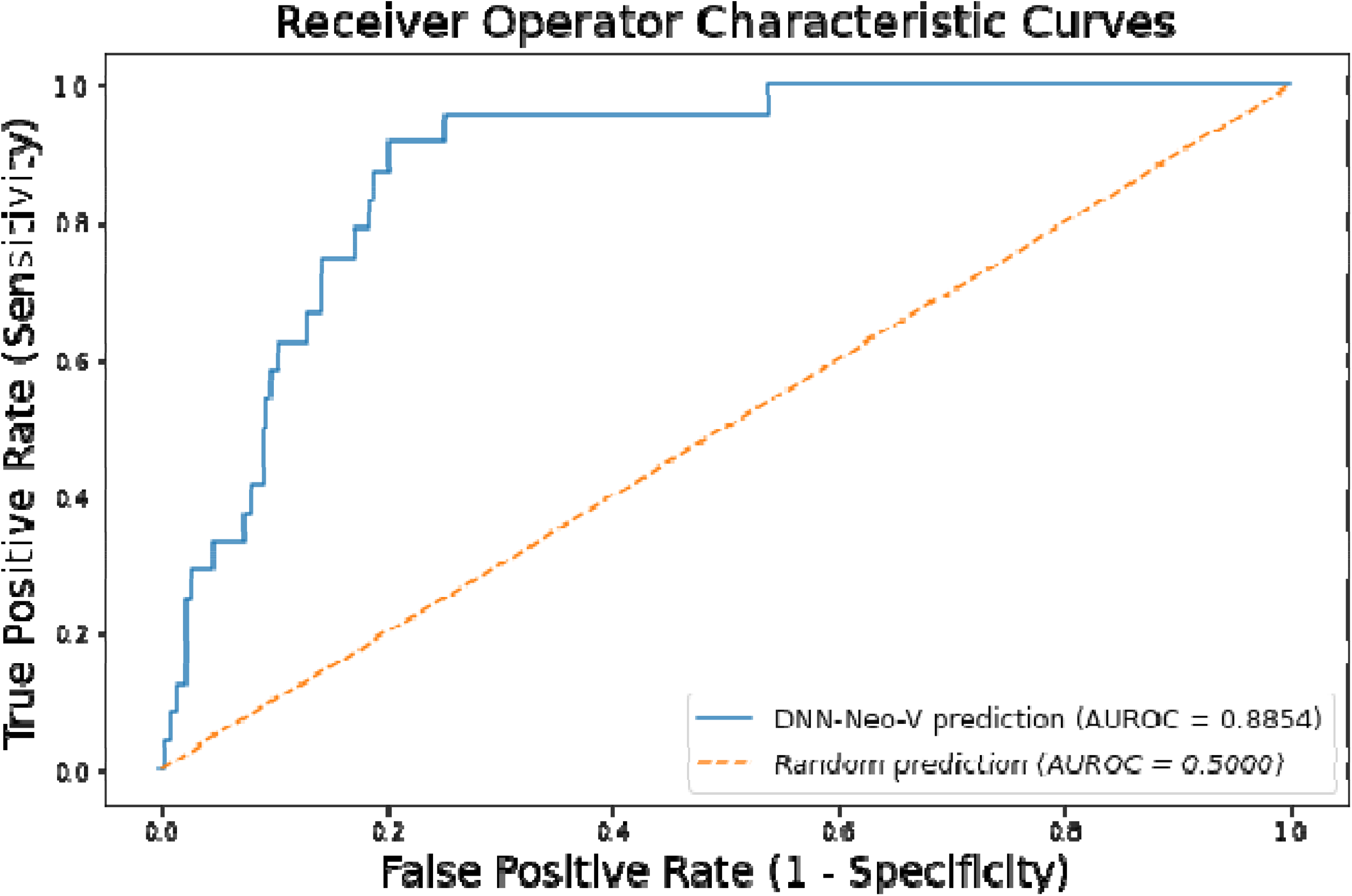
Receiver Operating Curve for Deep-Neo-V with AUROC

Machine learning has been used to predict mortality in the inpatient and the ICU setting in a number of different clinical conditions including our prior papers. The methods need to continue to evolve to have better outcome predictions, previously we developed the FLAIM-Framework approach, which was an attempt to build a workflow pipeline to produce more accurate results. The Neo-V Framework builds on our previous work and has the statistical power of FLAIM, but it can be used to apply deep learning to smaller datasets. We call this technique “horizontal expansion” of the dataset in which we horizontally expand the data by combining two or more variables to create new-variables (Neo-V). The combination was conditional that the variables were clinically relevant e.g. BUN and creatinine. In contrast, vertical expansion of the dataset is adding new patients.

We wanted to do a complete and thorough analysis of the Deep-Neo-V algorithm, so we compared it to other deep-learning models out there on COVID-19 mortality prediction currently available in medical literature(34). Our Deep-Neo-V model outperforms all our conventional models and our Deep-FLAIM model. It also outperformed the currently available Deep-learning model by Zhu J.S. *et al* in terms of training set accuracy, testing set accuracy, AUROC, Precision, specificity and positive predictive value(34). However, the Deep-Neo-V model underperformed in terms of sensitivity and slightly with negative predictive value. The Deep-Neo-V will continue to improve and develop and will potentially be replaced by a model with better performance parameters (accuracy, PPV and NPV). This model in its current configuration can be used to predict mortality after day-1 (considers labs and clinical characteristics in the first 24 hours) of hospital admission and can help in stratification of patients. It can help clinicians answer a number of questions and aid in decision-making like triaging patients, should an elderly patient receive aggressive treatment, or a younger patient receive life supportive management. These are tough questions and the model will give clinicians clarity about the course and plan for the patient. This model can also help clinicians with family counselling, appropriate decision making, limit excessive intervention or aggressive treatments and effective resource management. Like most digital tools this current algorithm is user friendly, can provide results instantaneously and easy to use. After further validation this model can be incorporated into hospital patient management systems and ready for clinical use.

The Deep-Neo-V model has some limitations in terms of the available dataset, retrospective nature of the dataset and data form a single hospital, analyzed at admission and day-one data, other observational study confounders may exist and are unaccounted for. In the immediate future we actively look to validate these findings in an external dataset. In the longer term we will continue to develop an algorithm built on the Neo-V Framework approach that has the potential to be implemented, initially in future pandemics because of its ability to accurately predict outcomes using smaller datasets.

## CONCLUSION

Deep-Neo-V is a statistically robust machine learning model that is developed for clinical use to predict mortality risk in patients admitted with RT-PCR proven COVID-19 infection. The mortality prediction was modeled based on clinically relevant variables (patient associated risk factors and the first 24-hours labs. Our experimental results show that with a high accuracy and specificity it has the potential to develop as a test of choice for predicting mortality in COVID-19 patients. These findings need further external validation.

## Data Availability

Available on request

## AUTHORS CONTRIBUTION

Maleeha Naseem: Study design, methodology, protocol design, IRB/ERC approval, data collection supervision, data collection tool and quality control, statistical analysis review, medical literature review and article writing.

Hajra Arshad: Data collection, protocol design, data collection tool, statistical analysis review, medical literature review and article writing.

Syeda Amrah Hashmi: Data collection, statistical analysis review, medical literature review and article writing.

Furqan Irfan. Medical literature review, statistical analysis review, and article writing.

Fahad Shabbir Ahmed. MD: Original concept, study design, methodology, data collection tool and construction of FLAIM, Deep-FLAIM and Neo-V frameworks, statistical analysis and review, python-coding for machine learning and article writing, editing, review.

---

**Supplemental Figure 1.**
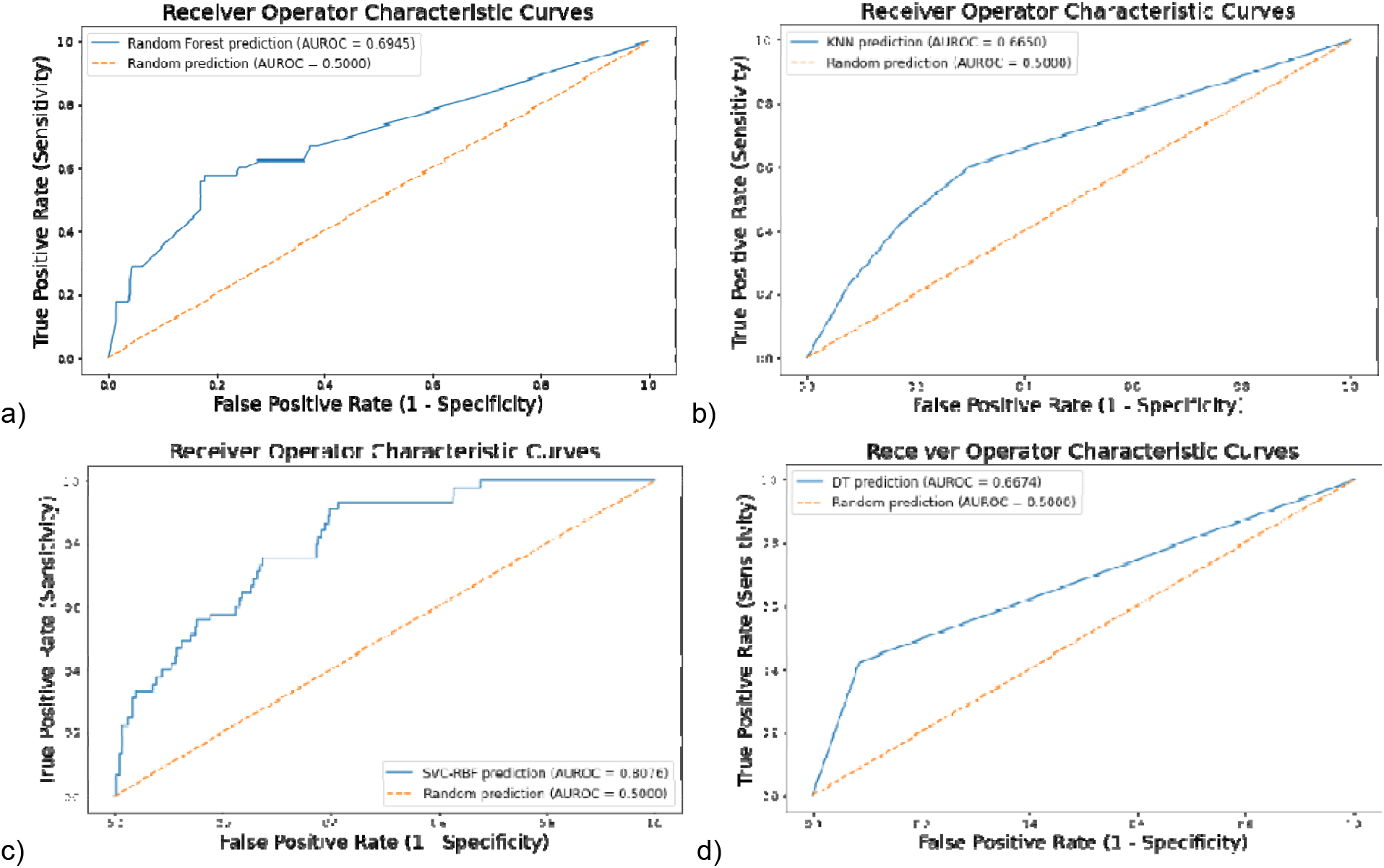
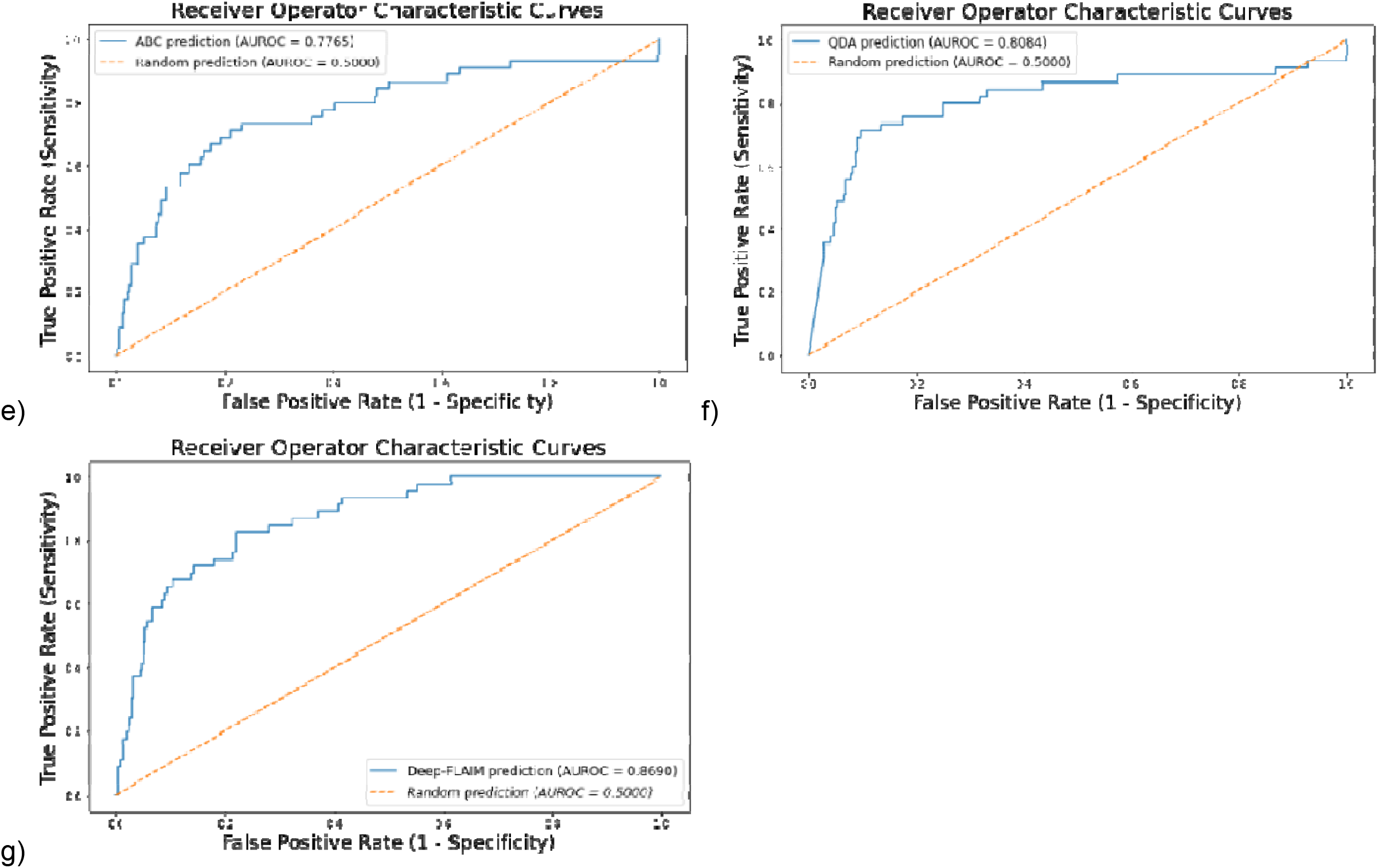
Receiver Operating Curve with AUROC for a) RF, b) kNN, c) SVC-RBF, d) DT, e) ABC, f) QDA and g) Deep-FLAIM

