## Supplemental Figure 1. for "Predicting mortality in SARS-COV-2 (COVID-19) positive patients in the inpatient setting using a Novel Deep Neural Network"

**Supplemental Figure 1**. Receiver Operating Curve with AUROC for a) RF, b) kNN, c) SVC-RBF, d) DT, e) ABC , f) QDA and g) Deep-FLAIM

a)
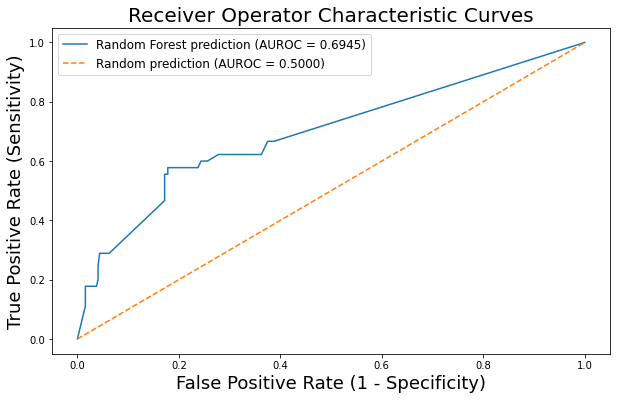
 b)
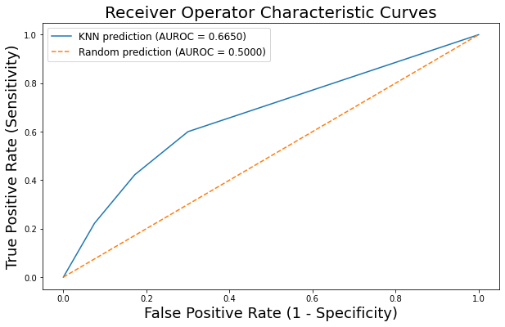

c)
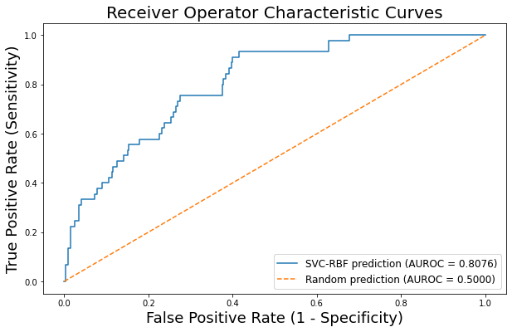
 d)
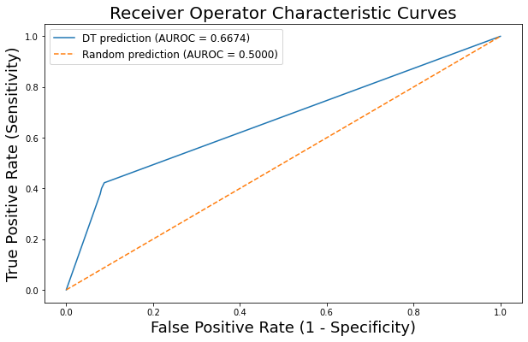

e)
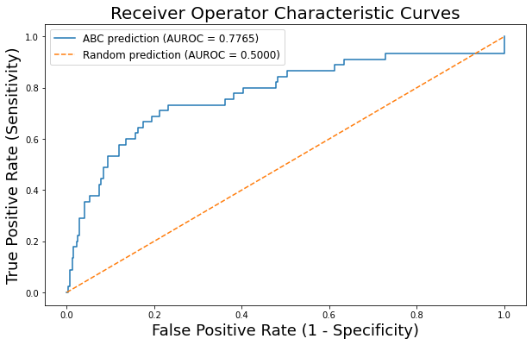
 f)
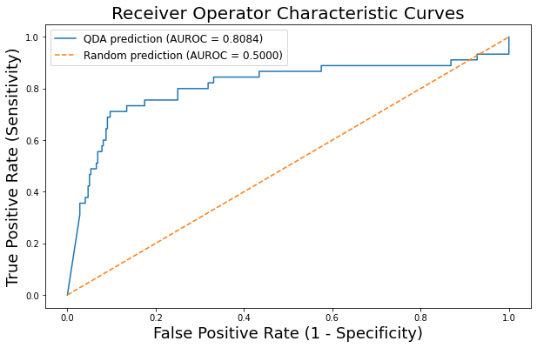

g)
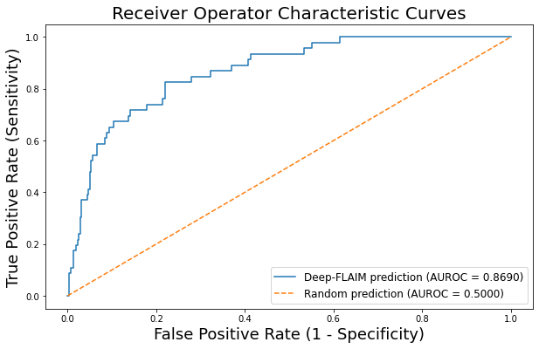
